# Dynamics of the Gut Microbiota in MEBO and PATM conditions: Protocol of a fully remote clinical study

**DOI:** 10.1101/2020.08.21.20179242

**Authors:** Irene S. Gabashvili

## Abstract

**Summary:** The proposed study will identify microbial communities associated with flare ups and remissions of MEBO (systemic malodor of metabolic origin) or PATM (“People allergic to me”) conditions.

**Background:** Human odor-prints, mostly owing to the microbiome, have proven their value as biomarkers of health and environmental exposures.

In recent years, microbial networks responsible for localized malodors (e.g., halitosis [1,2], groin area, foot and axillary odor [3,4]) have been mapped by using next generation sequencing approaches.

Intestinal microbes responsible for psychologically debilitating systemic malodor (whole-body and extraoral halitosis), however, remain to be identified. Even a relatively straightforward disorder of choline metabolism trimethylaminuria (TMAU) is thought to exhibit complex host-gene microbiome interactions [5] and has not been sufficiently studied.

Mapping gut microbiome is needed to understand human metabolic disfunction, make proper dietary recommendations and develop targeted treatments such as microbial therapies [6–8]. Our preliminary analysis of culture-, PCR- and 16S-RNA-based data found several Operational Taxonomic Units (OTUs) potentially linked to systemic malodor. Proposed controlled pilot study will provide a more comprehensive evaluation and, combined with our prior data [9–12], will help to develop new therapies and treatments.

## Aims

1. Use a longitudinal approach to determine if any characteristics of the microbiota are associated with episodes of metabolic malodor.
2. Examine the relationship between fecal microbiome composition, diet, and inflammatory/metabolic disease markers.

The hypothesis of this study is that, in spite of genetic heterogeneity, the pathology involves common patterns in the gut microbiome.

## Research Proposal

As randomized control trial (RCT) methodology is rarely used in community settings and is problematic when applied to supportive care in idiopathic malodor (smaller sample, unethical to randomize), proposed study is non-randomized.

To capture gut microbiota dynamics in idiopathic malodor, we will ask participants to continue their usual diet and collect a stool sample at the first opportunity after experiencing a flare-up (if condition is in active state) or a usual flare-up-free-day (if in remission). At the same time, subjects will be asked to submit observations of daily living, describing diet, quality of life, activities, stress and other environmental exposures (MEBO/PATM Life Quality Test and free-text notes).

We define a flareup as an episode of strong unpleasant odor emanating from the individual that can be detected at a social distance (4 feet or farther). In case of PATM (“People Allergic to Me” condition), the odor is less tangible but emanations cause constant allergy-like reactions from other people at social distances.

The first test will be followed by behavioral intervention for 8 weeks including personalized nutrition advice and education on stress management techniques.

Subjects will be asked to submit their second sample as soon as there is a (possibly temporary) improvement in their symptoms. Participants in remission or regression may choose to loosen their dietary restrictions to observe a minor flare-up and submit a sample at that time. If more kits are available, they will continue submitting more samples to better capture their physiological and psychosocial changes.

Microbiome data will be analyzed with software tools such as BCL2FASTQ, VSEARCH, Python libraries, diet analysis software [13], algorithms developed for analysis of noisy data [14–17], networks and metabolic pathways [18,19].

Application and the Protocol Narrative with all applicable attachments was submitted to MEBO Research Institutional Review Board (IRB) on March 6, 2018 and approved on May 11, 2018. Protocol Number: 201805110018MEBO

## Study Design and Timeline

### Study Type

Interventional (supportive intervention)

### Actual Enrollment

125 participants that volunteered to donate microbiome samples, including 112 participants interested in intervention, including receiving gut microbiome collection kits.

### Allocation

Non-Randomized

### Intervention Model

Sequential

### Masking

None (Open Label)

### Primary Purpose

Supportive Care; Prevention and Disease management

### Official Title

Dynamics of the Gut Microbiota in Idiopathic Malodor Production.

### Actual Study Start Date

June 16, 2018

### Actual Primary Completion Date

June 16, 2019

### Actual Study Completion Date

February 10, 2020

### Statistical Basis for sample size

No formal sample size calculation was performed. Previous studies of body odor found significant differences in axillar microbiomes of relatively small numbers of volunteers (50 times more bacteria in groups of 11 vs 13 Caucasians detectably different in axillary odor [20]). Simple estimates for a Wilcoxon test yielded similar sample sizes. Determining the minimum number of participants needed for microbiome trials is a largely unresolved subject because of excessive number of zeroes, over-dispersion, compositionality, intrinsically microbial correlations, and variable sequencing depths [21,

Kelly et al [23] found that for NOVA-type analyses five subjects per group in microbiome studies allows 90% power to detect a ω2 (adjusted coefficient of determination) of 0.05; 10 subjects per group allows 90% power to detect an ω2 of 0.02; and 20 subjects per group allows 90% power to detect an ω2 of 0.008. Omega-squared provides a less biased measure of effect size for ANOVA-type analyses by accounting for the mean-squared error.

Other studies examining differences in the gut microbiota in patients with suspected microbial imbalances in the usually included 30–60 patients or followed the rule of thumb of having at least 10 patients per event, to obtain sufficient statistical power for a reliable prediction [24

Obviously, larger sample numbers allow for higher accuracy. Our study is the largest ever remote trial on microbial basis of long-standing idiopathic malodor and PATM.

### Recruitment

Patients will be recruited from previous studies and non-interventional surveys. In addition, the study will be advertised on blogs, social media patient support groups and on https://Clinicaltrials.gov.

All participants are given links to the Quality of Life questionnaire and informed consent

(https://aurametrix.com/Studies/mebo-microbiome.html) prior to the beginning of the study and at each time point of sampling that will either be filled in by themselves or by a study coordinator via an online interview. Clinical data is collected by the study team from previously submitted health records and via a questionnaire. For stool sampling the participants will receive collection kits. Participants will collect gut microbiome samples using the kit instructions and mail it in the provided return envelope.

## Inclusion Criteria

- Idiopathic malodor or PATM symptoms experienced over a period of several months or years
- Able to read and understand the study information
- Willing and able to comply with questionnaires, nutritional recommendations, and other study procedures

## Exclusion Criteria

- Consistent inability/unwillingness to communicate symptoms and health-related concerns
- Consistent inability/unwillingness to distinguish physical symptoms from pure emotional reactions
- Lack of motivation to start feeling better
- Principal Investigator of this study, study coordinator and behavioral psychologist providing supportive care operate as a ‘virtual site’ as they did not physically work in the same location.

## Withdrawal of Subjects

Subjects can leave the study at any time for any reason if they wish to do so without any consequences.

## Self-Report Measures

Enrollment interview remotely administered by the study coordinator is asking about participants’ name, address for shipping test kits, age, sex, medical history, copies of relevant test results and diagnoses (TMAU urine test, genetic tests, halitosis, IBS, bromhidrosis, foot odor, vaginosis, gastroenterological tests, blood work, dermatological and dental evaluations, urological or endocrinologic disorders, etc.), detailed descriptions of odor conditions, ability to smell/detect malodor by themself, availability of a trusted buddy who could give them feedback about their odor or allergic reactions from others. Enrollees are informed that their personal information will be kept under strict privacy and confidentiality. Study coordinator will assign each human subject a de-identified unique identifier code. This ID will be used by subjects to submit their online survey responses and kit IDs that will be linked to medical histories. Research personnel will work with de-identified information.

Quality of Life (QoL) will be measured by the 24-item survey in addition to questions about IDs and informed consent checkbox. Quality of life assessment questionnaire is designed on the basis of the

Halitosis Associated Life-quality Test (HALT) and WHOQOL-100 questionnaires. Most questions were devised with a Likert scale of 0–5 where a higher score indicated a higher quality of life. Scores for five negatively framed questions are transformed to positively framed questions. MEBO test provides a total

QOL score (minimum score of 20 and maximum score of 150) and is focusing on four aspects of QOL: physical health, psychological health, social support and environment.

Electronic informed consent (eIC) contains all elements of informed consent required by HHS and/or

FDA regulations (45 CFR 46.116 and 21 CFR 50.25).

## Statistical analysis

Study coordinator will assign each human subject a de-identified unique identifier code. It will be used by subjects to submit their online survey responses and kit IDs that will be linked to medical histories.

Outcomes from questionnaires, tests and assays will be summarized for each group at different time points. The groups will be determined based on questionnaires and will include control group (individuals that never belonged to MEBO or PATM groups or are symptom-free relatives of study patients), subjects in remission, regression and active patients with symptoms of different severity. In addition

The data will then be examined for errors (e.g., out of range values), consistency, missing and spurious values. Missing data will not be imputed unless specifically noted. Any apparently spurious data will be verified. Non verified data will be excluded from summaries or analyses.

The first step in the data analysis will be an inspection for outliers and normality violations. Initial data exploration will be undertaken using inspection of frequency distributions and plots (i.e., graphical methods such as histograms and box plots), estimation of z-scores, estimation of skewness and kurtosis, and test for normality (i.e., analytic methods). If necessary, data will be transformed and centered for minimizing problems.

Descriptive statistics will be calculated and compared across all groups. Inter-group differences will be computed using statistical tests.

If the assumptions of the t-test cannot be met, yet the observations are independent, the Mann– Whitney test will be used for two group comparisons, if the data are at least ordinal in nature. Wilcoxon matched pairs test will be used for samples of the same subjects before and after improvement of symptoms.

Statistical analyses will be separately defined for each endpoint. There will be no adjustment of p-values for multiplicity unless specifically noted. P-values generated for this study are not intended to be conclusive, but provided for guidance only.

The composition of the microbiome will be measured in terms of both the number of unique types of microorganisms (i.e., α-diversity) and the microbial composition. We will calculate “phylogenetic entropy” to represent alpha diversity [26].

Microbiota composition will be evaluated in different ways. As microbiota data contain many variables, a first approach is to reduce the number of variables. Another approach will be longitudinal analyses with subjects acting as their own control. This will minimize the impact of confounding. We will also use other approaches for evaluation such as unsupervised clustering of microbiota profiles. Resulting clusters will be analyzed for over- or underrepresentation of different groups, again using the Mann-Whitney U test or the Student *t* test where appropriate.

Supervised machine learning methods will include regression and classification with different classifiers.Predictive performance of the model will be assessed by receiver operating characteristic (ROC) curves.

Using the above mentioned approaches, we can evaluate our data and decide which parameters can be used to create a model that can predict MEBO/PATM phenotype.

## Potential pitfalls and proposed resolutions

It is currently thought that 16S ribosomal RNA gene sequence analysis could not adequately uncover the potential TMA-producing potential and might not be suitable for systemic malodor research. However, if we don’t find significant statistical differences, we plan to apply resolution-enhancing methods similar to those developed by Nobel prize winner Joachim Frank for reconstruction of images from extremely noisy projections. We will also apply novel network analysis approaches. We are interested in functional profiling of gut microbes and identifying most prevalent metabolic pathways, as this will help to elucidate host physiological functions, in particular the underlying enzymatic deficiency.

The principal disadvantage of nonrandomized design is the potential for bias from confounding. The direction of this bias is unpredictable from study to study. For example, we may have a larger number of “sicker” patients in the intervention, thus biasing the trial against the intervention. Thus, we will evaluate our results in a larger context, and internal and external validity will be assessed through the replication of results in a variety of settings.

In the future, we’d like to see analysis extending beyond 16S RNA profiling because of the following:

A large subset of MEBO patients is convinced they are suffering from Candida overgrowth, as is often reinforced by naturopathic doctors. Some patients have had **yeast** infections of the mouth, scalp, skin, nails and genitals. There is also anecdotal evidence that anti-candida diet and antifungals may help to get rid of the odor. There were cases of **Blastocystis** spp. and **Giardia** infections. **Archaea** was shown to facilitate TMA reduction. In addition, almost half of MEBO patients have various skin conditions in which bacterial **viruses (bacteriophages)** are thought to play an important role.

## Results

The data is currently being analyzed. Final results are expected later this year

## Data Availability

Data and materials will be made available upon reasonable request.

https://aurametrix.com/Studies/mebo-microbiome.html

